# Neuroscience-Informed Psychoeducation for Recovery (NIPER): A Program to Promote Metacognition in People with Substance Use Disorders

**DOI:** 10.1101/2020.06.22.20137463

**Authors:** Tara Rezapour, Mohammad Barzegari, Elham Sharifi, Nastaran Malmir, Hamid Reza Ghiasvand, Mohammad Salehi, Alireza Noroozi, Hamed Ekhtiari

## Abstract

**Background:** A brief neuroscience-informed psychoeducation program (neuroscience-informed psychoeducation for recovery, NIPER) was developed to promote awareness (metacognition) in the main cognitive domains affected by drug and alcohol use to increase willingness to invest time and effort in the brain and cognition recovery process. The primary aim of this study was to determine the feasibility and acceptability of the NIPER program and its potential effectiveness in terms of increasing metacognition, psychological wellbeing and willingness for the brain and cognition recovery programs among people with substance use disorders (SUDs).

**Methodology:** 56 individuals with SUDs recruited from four outpatient treatment centres in Tehran, Iran and attended four 90-min sessions delivered weekly adjunct to their treatment as usual. The effectiveness was measured in terms of metacognition, and psychological wellbeing at baseline and at the end of the program. Rate of adherence and participation as well as willingness to continue with brain and cognition recovery programs were measured as feasibility outcomes.

**Results:** A total of 51 participants completed the study. Compared to the baseline assessments, participants reported significantly higher problems in dimensions of attention, memory, inhibitory control, decision making, motor/speech, interocpetion and insight, as well as higher level of psychological wellbeing (t=4.66. p<0.001). In terms of feasibility outcomes, the adherence and participation rates were found above 85%. The majority of participants expressed their high willingness to continue the brain and cognition recovery programs (86.2%) and introduce NIPER to their peers (98%).

**Conclusion:** Taking into account the results in terms of feasibility and preliminary effectiveness of NIPER in clinical context of addiction treatment, we consider NIPER as a potentially beneficial interventions to be offered to people with SUD to increase their awareness and engage them to the brain and cognition recovery process. The clinical efficacy of the intervention should be tested in future randomized clinical trials.

## Introduction

As a brain disorder, addiction is characterized by a broad range of apparent and subtle cognitive deficits including attention, episodic memory, executive functions (e.g., inhibitory control, flexibility, planning)(Rezapour, 2020; A. Verdejo-Garcia, Lorenzetti, V, Manning, V, Piercy, H, Bruno, R, Hester, R, Pennington, D, Tolomeo, S, Arunogiri, S, Bates, ME, Bowden-Jones, H, Campanella, S, Daughters, SB, Kouimtsidis, C, Lubman, DI, Meyerhoff, DJ, Ralph, A, Rezapour, T, Tavakoli, H, Zare-Bidoky, M, Zilverstand, A, Steele, D, Moeller SJ, Paulus, M, Baldacchino, A, Ekhtiari H., 2019). These deficits in substance users are clinically important, as they may contribute to poor treatment outcomes indicated by high risk of dropout, low treatment compliance, and shorter abstinence (Bruijnen et al., 2019). Moreover, cognitive deficits may affect individual’s self-efficacy and interfere with psychosocial, occupational and daily living functioning (Bruijnen et al., 2019; Weber et al., 2012).

Studies considering cognitive functions in substance users reveals that chronic use of drugs and alcohol may also negatively affect another component of cognitive functions termed as awareness or metacognition (Balconi, Finocchiaro, & Campanella, 2014). Metacognition is defined as individual’s ability to understand his/her own cognitive functions and use this understanding to regulate them (Balconi et al., 2014; Wasmuth, 2015). Impairment of this ability has been reported in previous studies among substance users using neuroimaging, self-reports, and behavioral measurements (Goldstein et al., 2009; Jung, Kim, Kim, Oh, & Kim, 2011; Maremmani et al., 2012; Williams, Olfson, & Galanter, 2015). For example, it has been shown that functional and structural alteration in regions such as rostral anterior cingulate, anterior insula and precuneus in people with SUDs are linked to the lack of self-awareness. Behavioral and self-report measures also indicated discordances between individual’s self-assessments and their actual performance on cognitive tasks (Moeller et al., 2010) or their informant report on the existence of cognitive problems in people with SUDs (A. Verdejo-Garcia & Perez-Garcia, 2008).

These experimental results of metacognitive deficits become increasingly important, when they contribute to the lack of insight and affect treatment outcomes. Substance users with poor metacognition are more reluctant to initiate or continue treatment and more probable to underestimate the need for changing behavior (Dean, Kohno, Morales, Ghahremani, & London, 2015).Thus improvement in metacognition may remove motivational barriers to invest time and effort in the brain and recovery process and improve treatment outcome. Despite the importance of metacognition in the recovery process in substance users, there is lack of intervention to target this function.

The neuroscience-informed psychoeducation for recovery (NIPER) program is designed as the first package in the field of drug addiction to raise individual’s awareness about cognitive deficits (metacognition) associated with using drug and alcohol as well as to motivate substance users to invest time and effort for their brain and cognition recovery process (Ekhtiari, Rezapour, Aupperle, & Paulus, 2017). In order to determine whether the intervention is feasible to be delivered to people with SUDs who receiving treatment as usual, we conducted a one-arm open-label trial in people with SUDs recruited from outpatient treatment centers. We assumed that providing individual with NIPER, may lead to improve their metacognition and increase their willingness to invest time and effort on the brain and cognition recovery programs.

## 2. Methods

### 2.1. Study setting

This is a single arm, four centers trial conducted to assess the feasibility of a psychoeducation program designed for people with SUD carried out in Tehran, Iran between July and October 2019.

To identify the interested outpatient centers, we advertised on the social media channels related to drug and alcohol addiction professionals and provided information about the content and length of program. After initial expression of interest to participate, 4 centers (one academic and three private centers) agreed to take part in this study by providing 56 volunteer participants. From these participants, 16 were recruited from center A, 17 were recruited from center B, 8 were recruited from center C, and 15 were recruited from center D and trained in groups up to 15. As a total we had six groups (two groups from center A, two groups from center B, one group from center C, and one group from center D). In these centers various models of addiction treatment including opioid agonist pharmacotherapies with methadone, buprenorphine and opium tincture were provided for people with opioids use disorder, as well as intensive outpatient psychosocial interventions for treatment of stimulants use disorder.

### 2.2. Participants

Since this study is a feasibility study, the sample size was not calculated and it was determined about 56 participants, based on the guidelines suggested for feasibility studies (Billingham SA, 2013). We recruited participants who (1) were medically stable; (2) were able to speak and write in Farsi; (3) were aged between 18-65 years; (4) were diagnosed with opioid and/or stimulant use disorders based on the Diagnostic and Statistical Manual of Mental Disorders (DSM–5); (5) had received any standard treatment program (STP) based on the Iran Ministry of Health protocols and guidelines for more than 2 weeks and less than 24 weeks; and (6) were willing to participate in the research. Individuals who (1) were unable to perform assessments and comprehend intervention-related information; (2) were concurrently participating in another study receiving similar types of intervention beside STP which might interfere with our program; (3) had major uncontrolled psychiatric disorders (including depression, bipolar or psychotic disorders); and (4) had a history of suicidal attempt during last year, were excluded from the study. Eligibility criteria were assessed using self-report data. All the included participants signed the written informed consent after they were provided by all the necessary research-related information. Those participants who remained in the program and completed pre and post assessments were compensated at the end of the study for the time they spent in research and received a certificate of course completion. This study has been approved by the Tehran University of Medical Sciences research ethics committee (IR.TUMS.VCR.REC.1398.771).

### 2.3. Intervention

The intervention used in the present study was the neuroscience-informed psychoeducation for recovery (NIPER) program developed to promote individual’s metacognition as well as compensatory strategies and healthy life-style which may support brain healing during addiction recovery (Ekhtiari et al., 2017). NIPER is a paper-based program consisting of four group sessions, that each session is estimated to last 90 minutes (two 45-min parts, separated by a 10-min break). NIPER translates knowledge from neuroscience of addiction into individual’s everyday life within three modules:

#### Part I: Brain Literacy Module

“What did drug/alcohol do to my brain? What are the signs of brain deficits caused by drug/alcohol? How do you experience these brain deficits in everyday activities?”. These are the main questions that may be raised by people with SUD who are planning for their recovery. To answer these questions, NIPER applies the Addictions Neuroclinical Assessment (ANA)(Kwako, Momenan, Litten, Koob, & Goldman, 2016) and Research Domain Criteria (RDoC)(Insel T, 2010) frameworks to define the most affected brain functions by using drug/alcohol. According to these frameworks, people with SUD experience a various profile of impairments in negative valence, positive valence, cognitive control, attention, memory, perception and understanding of self/others, arousal and motor functions. Each system is composed of different subcomponents that are depicted as the circles in the Figure 1.

**Figure 1.**
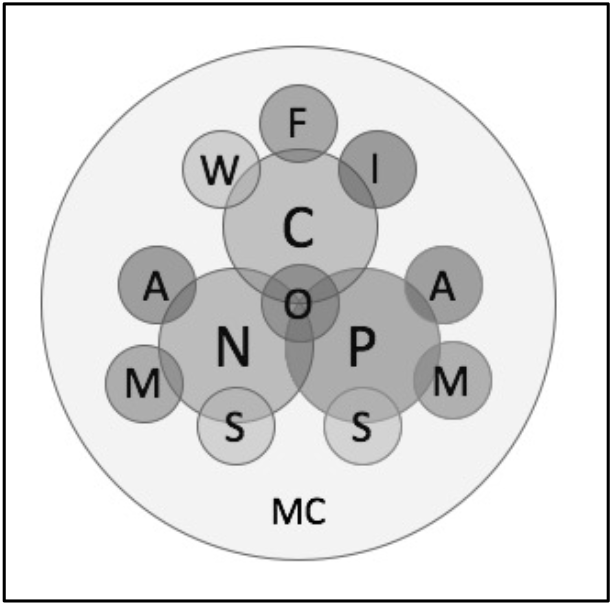
Main cognitive domains addressed in the neuroscience-informed psychoeducation program for recovery (NIPER). The three main cognitive systems (bigger circles) include: N= negative valence, P= positive valence, C= cognitive control. The subcomponents are working memory (W), saliency processing (S), inhibition (I), flexibility (F), interoception (O), attention (A), memory (M), metacognition (MC) to self (insight) and others (social cognition).

Each session starts with a first-person narrative of a subjective experience of cognitive deficits associated with drug and alcohol use and colorful cartoons depicting the relevant sign emerges in real life. Table 1 depicts all the cartoons used for this module as well as the related scenario for each main domains of cognitive deficits. Brain literacy modules encourage individuals to share their personal experiences of having similar cognitive deficits and their associated problems in their daily lives with others in the group. After group discussion, participants are provided by a series of paper-pencil exercises that demand using the discussed cognitive functions (e.g., “spot the difference” exercise for attention and the different stroop exercises for inhibitory control). By practicing this part, individuals gain a deeper understanding of the of the discussed cognitive functions. This module takes around half of the time in each session.

**Table 1.**
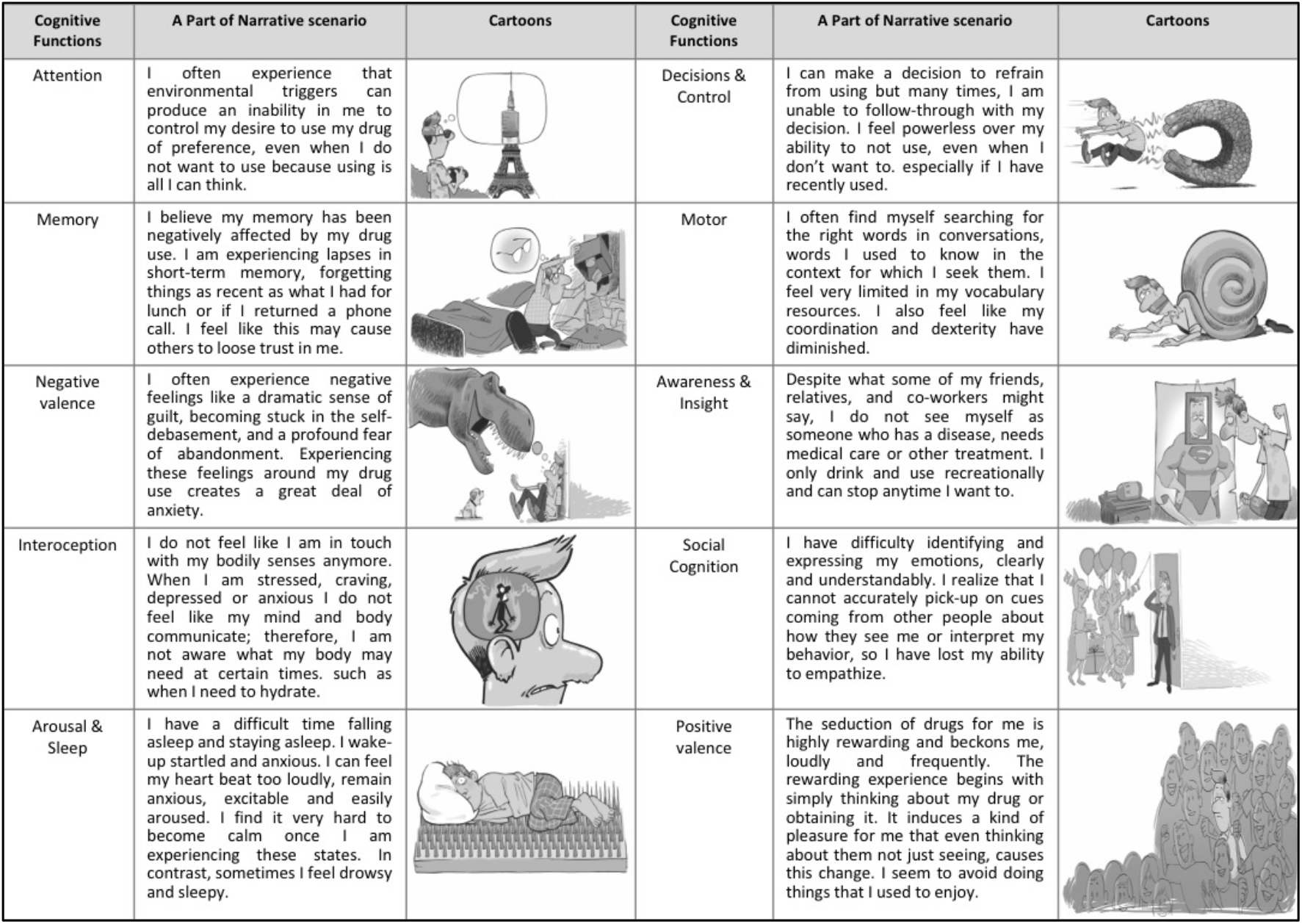
The brain literacy modules of NIPER including cognitive domain, narrative scenario and cartoons designed in 10 domains and distributed in 4 sessions.

| Cognitive Functions | A Part of Narrative scenario | Cartoons | Cognitive Functions | A Part of Narrative scenario | Cartoons |
| --- | --- | --- | --- | --- | --- |
| Attention | I often experience that environmental triggers can produce an inability in me to control my desire to use my drug of preference, even when I do not want to use because using is all I can think. |  | Decisions & Control | I can make a decision to refrain from using but many times, I am unable to follow-through with my decision. I feel powerless over my ability to not use, even when I don't want to, especially if I have recently used. |  |
| Memory | I believe my memory has been negatively affected by my drug use. I am experiencing lapses in short-term memory, forgetting things as recent as what I had for lunch or if I returned a phone call. I feel like this may cause others to lose trust in me. |  | Motor | I often find myself searching for the right words in conversations, words I used to know in the context for which I seek them. I feel very limited in my vocabulary resources. I also feel like my coordination and dexterity have diminished. |  |
| Negative valence | I often experience negative feelings like a dramatic sense of guilt, becoming stuck in the self-debasement, and a profound fear of abandonment. Experiencing these feelings around my drug use creates a great deal of anxiety. |  | Awareness & Insight | Despite what some of my friends, relatives, and co-workers might say, I do not see myself as someone who has a disease, needs medical care or other treatment. I only drink and use recreationally and can stop anytime I want to. |  |
| Interoception | I do not feel like I am in touch with my bodily senses anymore. When I am stressed, craving, depressed or anxious I do not feel like my mind and body communicate; therefore, I am not aware what my body may need at certain times. such as when I need to hydrate. |  | Social Cognition | I have difficulty identifying and expressing my emotions, clearly and understandably. I realize that I cannot accurately pick-up on cues coming from other people about how they see me or interpret my behavior, so I have lost my ability to empathize. |  |
| Arousal & Sleep | I have a difficult time falling asleep and staying asleep. I wake-up startled and anxious. I can feel my heart beat too loudly, remain anxious, excitable and easily aroused. I find it very hard to become calm once I am experiencing these states. In contrast, sometimes I feel drowsy and sleepy. |  | Positive valence | The seduction of drugs for me is highly rewarding and beckons me, loudly and frequently. The rewarding experience begins with simply thinking about my drug or obtaining it. It induces a kind of pleasure for me that even thinking about them not just seeing, causes this change. I seem to avoid doing things that I used to enjoy. |  |

#### Part II: Brain Recovery-Supporting Activities Module

The second half of each session starts with providing ideas on the recovery supporting activities that engage the cognitive functions addressed in the brain literacy module. For this module, patients are asked to practice these activities regularly and talk about their personal experiences in the next session. These activities can be easily embedded in daily life activities and do not need any particular tools. For example, to reinforce memory function, participants are trained to record daily events in a notebook, named as brain book and try to visualize events as they record. These activities are selected from routine exercises practiced in cognitive rehabilitation programs.

#### Part III: Healthy Brain Lifestyle Module

Each session ends with triggering the curiosity of individuals by asking this question that “What can we do to help the process of brain healing?”. Prior to initiate this module, it is very important to explain participants about the length and speed of brain healing process during addiction recovery. NIPER uses “a broken hand” metaphor for the brain in the recovery process to explain how brain needs active support to gradually obtain the normal cognitive functions in the process of recovery (similar to range of motion for broken hand). A set of evidence-based recommendations which may foster brain healing process, are provided in the form of to-do’s to improve recovery-supporting lifestyle. These recommendations emphasize on the components such as healthy diet, physical activities, social communications, mental activities, and sleep quality and their importance for the brain recovery.

Therefore, NIPER provide people with SUD with critical information about brain functions that are affected by drug and alcohol use as well as activities and strategies that may promote brain healing process during the course of addiction treatment. NIPER has been developed to help people with SUD to know more about addiction as a brain disease and use this awareness in their real life. The content of the three NIPER modules is described in more detail in Table 2. NIPER has been originally developed in grayscale version and consists of a 116 A-5 size booklet.

**Table 2.** An overview to the content of the three modules in the NIPER program during 4 sessions.

| Session | Brain Literacy Module | Brain Activities Module | Healthy Lifestyle Module |
| --- | --- | --- | --- |
| First | 1. Attention<br>2. Memory | 1. Do Word Exercises<br>2. Be Your "Present-Moment" Attention Coach<br>3. Train Your Brain to Be Flexible<br>4. Journal in Your Brain Book<br>5. Play "Memory Games"<br>6. Reduce "Brain Clutter" | 1. Be More Mentally Active<br>2. Be a Healthy Foodie |
| Second | 1. Negative Valence<br>2. Interoception<br>3. Arousal & Sleep | 1. Use Positive Language<br>2. Live in Gratitude<br>3. Volunteer for Charity Work<br>4. Practice Body-Presence<br>5. Observe Your Heart Rates<br>6. Practice Mindfulness<br>7. Create a Sleep Heaven<br>8. Pamper Yourself Occasionally<br>9. Enjoy the Benefits of Warmth/Heat | 1. Be Calm and Relaxed<br>2. Be in Tune with Your Emotions<br>3. Be a Healthy Sleeper |
| Third | 1. Decision & Control<br>2. Motor | 1. Set Daily Goals<br>2. Track your Money<br>3. Practice Patience<br>4. Practice Paraphrasing<br>5. Enjoy the "Artist in You"<br>6. Improve Dexterity | 1. Commit to Abstinence from Intoxicants<br>2. Be More Physically Active |
| Forth | 1. Positive Valence<br>2. Social Cognition<br>3. Self-Awareness | 1. Observe Your Brain Processes<br>2. Attend to Your Posture<br>3. Live Weight-Conscious<br>4. Use Compassion and Understanding<br>5. Allow Yourself to be Transparent<br>6. Be a Voice Analyzer<br>7. Be a Member of the Happiness Club<br>8. Be a "Hobbyist"<br>9. Detox Your Brain from Negative Memories by Making Positive Ones | 1. Be a Healthy Friend to Yourself<br>2. Be a More Socially Active<br>3. Be Patient and Hopeful |

It should be noted that in this study, NIPER was delivered by four trained providers (MB, ES, NM, HG), each communicated with one clinical site constantly during the study. All the providers had at least a bachelor’s degree in psychological or social sciences and prior clinical experiences working with people with SUD. To improve treatment fidelity, providers had been trained about the intervention through several online meetings before this study and sessions were weekly supervised by the program developers (HE and TR). They were also asked to record the exact amount of time spent for each training session and reported the extra time if they spent for participants individually. Moreover, to ensure that the training materials were delivered equally for all the training groups, NIPER was offered with a structured booklet, given similarly to all training groups (Bellg et al., 2004).

### 2.4. Measures

Training providers collected self-report data from participants at baseline (week 0) and at the end of intervention (week 4) including basic information, metacognition, and psychological wellbeing. Data about feasibility outcomes were also collected in terms of adherence and participation rate as well as participants’ opinions about the program.

#### 2.4.1. Patients basic information

Basic information including sociodemographic (age, years of education) and addiction related data (main substance use, age of onset, years of use, previous addiction treatment experience) were collected at baseline.

#### 2.4.2. Metacognition

Metacognition was evaluated using a developed instrument based on the NIPER’s cognitive domains inspired by the Measure of Insight into Cognition (Saperstein, Thysen, & Medalia, 2012). This instrument includes 10 self-report items asking individuals whether they have perceived cognitive problems rating on a 5-point Likert scale (0=not at all to 5= a lot). The items are adjusted to tailor cognitive functions in the NIPER program consisting of difficulty with attention and concentration, declined in memory function, diminished in behavioral control, difficulties in decision making, difficulties in speech and movement, distorted brain-body connection, sleep problems, increased negative emotions, decreased positive emotions, difficulties in social interactions, and declined in self-awareness and insight. Higher score for each item, represent higher level of perceived impairments. The participants were asked to fill out the instrument at baseline as well as at the end of the intervention.

#### 2.4.3. Psychological wellbeing

To assess the effect of program on the psychological wellbeing, we used psychological subset of the World Health Organization Quality-of-Life Scale (WHOQOL-BREF) (Nejat et al.,2006).This subset includes six items that focuses on the ability to concentrate, self-esteem, body image, spirituality (i.e. the extent to which they feel their life is meaningful), and the frequency of positive or negative feelings (i.e. blue mood, despair, anxiety, depression). Participants were asked to rate each item on a 5-point scale (0= not at all to 5= extremely amount). The total score was then transformed on a 0–100 scale, in which higher scores indicated higher psychological wellbeing. In the present study, we used the Persian version of the scale validated by (Karimlou M, 2011).

#### 2.3.4. Feasibility outcomes

To collect data about the feasibility of the program, rate of adherence and participation were evaluated by the number of participants who remained in the intervention (who completed pre and post assessments and attended to least 3 from four sessions) and the number of participants attended each session, respectively. They were also asked to rate their willingness to introduce the NIPER to their peers as well as to continue investing time and effort in the brain and cognition recovery process (e.g., cognitive training/rehabilitation). These questions were asked after the intervention.

## 3. Results

### 3.1. Sample characteristics

Of the 56 individuals who participated in the NIPER intervention and completed the baseline assessments, 51 remained and re-assessed at the end of the intervention. 5 participants dropped out after the first session due to uncertain reasons. All participants were men and were recruited during their early phase of abstinence (first month of abstinence). Table 3 indicates the descriptive characteristics of sample.

**Table 3.** Demographic and substance use related data of the retained sample (n=51)

|  | Mean (SD)/N (%) |
| --- | --- |
| Age | 38.04(9.97) |
| Years of education | 11.45(3.34) |
| Days of abstinence | 23.91(10.3) |
| Main substance used |  |
| Opioids | 40(78.4%) |
| Stimulants | 11(21.5%) |
| Age of main substance use onset | 22.94(6.54) |
| Years of main substance use | 9.86(6.15) |
| Previous addiction treatment experiences |  |
| Yes | 42(82.3) |
| No | 8(17.6) |
| History of injection |  |
| Yes | 6 (11.8) |
| No | 44 (86.3) |

### 3.2. Metacognition

To compare metacognition from baseline to post-intervention, two paired-sample t-tests was used for each item. As shown in Table 4, participants reported significantly higher impairments in the domains of attention, memory, control and decision making, motor and speech, interoception and insight. While the mean scores of participants increased for the other dimensions, they were not statistically meaningful (p>0.05).

**Table 4.**
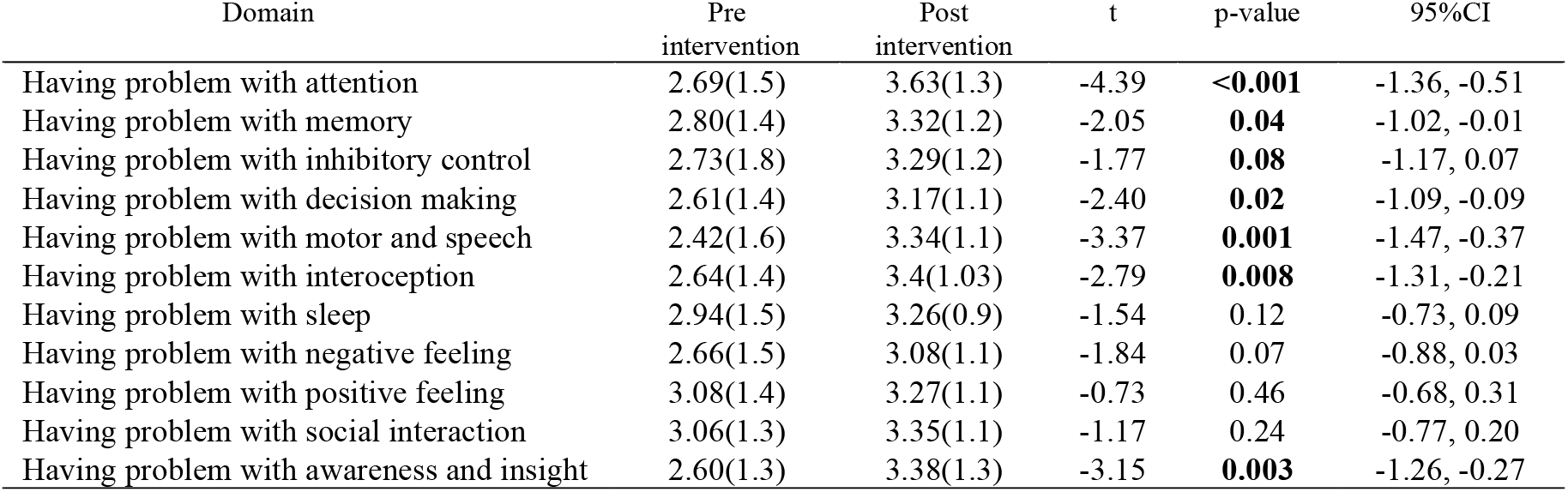
Changes in perceived cognitive impairments from baseline to post intervention (n=51)

### 3.3. Psychological wellbeing

At baseline, the mean psychological wellbeing score was 36.14 (SD=15.17) and at the end of intervention, this score increased to 43.39 (SD=16.01), indicating a significant improvement (t(50)=-4.66, p<0.001, 95% CI [−10.37, −4.13]).

### 3.4. Feasibility outcomes

The adherence rate was 91% (51 from 56 participants completed the intervention and pre-post assessments). The participation rate was calculated for each session in terms of the total number of participants who were attended. The results were as follows: 96% (n=49) for the first session, 94% (n=48) for the second session, 96% (n=49) for the third session, and 98% (n=50) for the fourth session. Regarding the willingness to continue with brain and cognition recovery programs, one participant (2%) rated his response as very low, six participants (11.8%) rated as medium, seventeen participants (33.3%) rated as high and twenty-seven (52.9%) rated as very high. Finally, the majority of participants (n=50, 98%) said that they would introduce the NIPER to their peers.

## 4. Discussion

This study aimed to test the feasibility of a new neuroscience-informed psychoeducation program in the context of addiction recovery. The results indicated that the intervention is feasible and acceptable to be implemented for people with SUD with potentials to improve their understanding about their deficits related to attention, memory, behavioral control, decision making, motor and speech, interoception, and insight. These results should be interpreted cautiously, since we we didn’t have a control group and these changes could also be related to the progress in the standard treatment program during the one month of the intervention. Regarding with feasibility measures, we found an acceptable rate of adherence and participation over the course of intervention as well as high rate of willingness to continue it.

We didn’t find any significant difference between individual’s self-report on sleep, negative feeling, positive feeling and social interaction problems between pre and post assessments. These deficits might be more apparent to the individual to perceive and detect in the everyday functioning even before receiving the NIPER.

NIPER is among the first attempts to translate the brain-related topics in the field of addiction into a standalone psychoeducational program with therapeutic intention. In parallel to our study, similar neuroscience-based approaches were used to develop prevention program to reduce the risky use of drug and alcohol in young students through providing them with neuroscience-based information on addiction (Debenham, Birrell, Champion, Askovic, & Newton, 2020).

Following this feasibility study, further studies are necessary in order to conclude about the efficacy of the NIPER in addiction medicine. Future studies should be designed in forms of randomized clinical trial with larger sample size and control groups. These trials should consider the impact on the clinical outcomes and monitor potential changes over a follow-up period; however, the expectations should not exceed the potentials for a 4 sessions brief intervention.

The impact of improved metacognition on recovery process has been broadly investigated in different psychiatric disorders (e.g., schizophrenia, bipolar disorder). This relation has been investigated through measures including medication acceptance and therapeutic alliance (Moritz et al., 2018). Improved metacognition may also affect outcomes of brain and cognition recovery programs (e.g., cognitive rehabilitation) in two ways. First, people with SUD who acquire this awareness may perceive these interventions as more meaningful and necessary for the addition recovery process. Thus, they become more motivated to actively participate in the cognitive training/rehabilitation programs and less likely to drop out which is a challenging part in the addiction treatments. Secondly, this neuroscience-informed look towards addiction and its brain deficits and potentials for the brain recovery may be beneficial for those individuals who deny their current deficits and are not hopeful for the brain recovery even when they are aware of these brain deficits. This new metacognitive awareness may reduce resistance to the therapeutic interventions.

In conclusion, this pilot study demonstrates the feasibility and acceptability of implementing a neuroscience-informed psychoeducational intervention for substance users. We are hoping that providing such educational program for people with SUD who are commonly suffer from lack of proper understanding about their cognitive deficits, could enable them to recognize their problems, assign them to their drug use disorder and perceive the need for appropriate treatment. Offering these types of metacognition enhancing programs, may be even more crucial at the early stage of addiction treatment (Maremmani et al., 2012) once individuals are more uninformed and ignorant about their addiction related problems particularly the cognitive ones.

## Data Availability

The study is a feasibility study.

## Acknowledgements

We gratefully acknowledge the contribution of Iranian National Center for Addiction Studies (Mehri Nouri, Amir Azarbara, Hassan Ghafouri), Etemad (Dr. Behbood Aghazadeh), Mehr Aeen (Dr. Mohammad Salehi), and Malekzadeh clinics (Dr. Ali Malekzadeh) who were participated in this study. Authors thank Dr. Martin Paulus and Mr. Brad Collins for their contribution in the development of the NIPER materials.

## Authors’ Contributions

HE, AN and TR designed the study. MB, ES, NM and HG run training sessions and collected data and contributed in the initial data analysis. HE and TR developed the initial draft of the manuscript and performed the statistical analysis. All authors read, revised and approved the final draft of the manuscript.

